# Comparison of gene expression in living and postmortem human brain

**DOI:** 10.1101/2023.11.08.23298172

**Authors:** Leonardo Collado-Torres, Lambertus Klei, Chunyu Liu, Joel E. Kleinman, Thomas M. Hyde, Daniel H. Geschwind, Michael J. Gandal, Bernie Devlin, Daniel R. Weinberger

## Abstract

Molecular mechanisms of neuropsychiatric disorders are challenging to study in human brain. For decades, the preferred model has been to study postmortem human brain samples despite the limitations they entail. A recent study generated RNA sequencing data from biopsies of prefrontal cortex from living patients with Parkinson’s Disease and compared gene expression to postmortem tissue samples, from which they found vast differences between the two. This led the authors to question the utility of postmortem human brain studies. Through re-analysis of the same data, we unexpectedly found that the living brain tissue samples were of much lower quality than the postmortem samples across multiple standard metrics. We also performed simulations that illustrate the effects of ignoring RNA degradation in differential gene expression analyses, showing the effects can be substantial and of similar magnitude to what the authors find. For these reasons, we believe the authors’ conclusions are unjustified. To the contrary, while opportunities to study gene expression in the living brain are welcome, evidence that this eclipses the value of postmortem analyses is not apparent.

## Introduction

Studies of gene expression in postmortem human brain have become an important research approach to translate genetic associations with neuropsychiatric illness into molecular associations of disease mechanisms. Yet, questions have been raised about the fidelity of postmortem tissue as representative of disease biology in the living brain. By contrasting transcriptomes from living (LIV) and postmortem (PM) tissue samples, Liharska and colleagues report that ≈80% of the gene level transcripts are differentially expressed (DE) at FDR < 0.05 (Liharska et al. 2023), implying that all or virtually all of these transcripts differ in their abundance when LIV and PM transcriptomes are contrasted. These are vast differences, as Liharska and colleagues note, from which they reach two bold conclusions: (1) postmortem tissue may not be a good proxy for living tissue in human brain research; and (2), molecular signatures identified in postmortem human brain samples may not be accurate representations of disease processes occurring in the living brain. However, neither of these conclusions are actually supported by their underlying data, and both are in direct conflict with a wealth of published literature suggesting otherwise. Furthermore, as we demonstrate in detail below, there are serious unaddressed statistical confounds in the data and analyses presented in Liharska and colleagues, which make any conclusion from this study entirely premature. In particular, and surprisingly, the quality of the RNAseq data from the living tissue of Liharska et al is inferior in most respects to the PM tissue in their report.

Regarding (1), experienced researchers would not assume postmortem tissue is a perfect proxy for living tissue. To the contrary, it is well known that gene expression patterns in post-mortem tissue differ from those of living tissue, even in perfectly controlled experimental conditions (Catts et al. 2005, Pozhitkov et al. 2017, Ferreira et al. 2018, Heng et al. 2021). Shortly after death, a portion of the RNA transcripts begin to degrade. Their absolute abundance declines with increasing post-mortem interval – predictably, both in terms of the rate of decline and the nature of the transcripts showing decline. A smaller fraction increases after death, again predictably. Nonetheless, most RNA transcripts show no detectable change in their absolute abundance over postmortem intervals (PMI) typical of human research. We use the term “absolute”, as in “absolute abundance”. We do so because Liharska and colleagues employ a commonly used procedure for quantification of RNA transcripts, RNA sequencing, that characterizes the *relative abundance* of different transcripts. The sequencing approach they employ targets a fixed number of RNA sequence reads per sample, which are determined by the sequencing run and distributed among different transcripts according to their relative frequencies. Each gene’s transcript expression per subject is then quantified as counts per million sequence reads or CPM. Different experimental or treatment conditions between the LIV and PM tissues, together with quantification via CPM, is sufficient to cause vast differences in CPM across genes between these tissue types, even when the absolute abundances of most transcripts per gene are unchanged. Normalization methods such as those implemented in edgeR attempt to remove this composition bias (Robinson and Oshlack, 2010). In simulation analyses (see Results below), we provide one such example, involving the well-known phenomenon of RNA degradation for a subset of transcripts. Importantly, if this were the sole concern, it would be easily rectified, as we also show. We also re-analyze the dataset by Liharska and colleagues (henceforth Liharska) using qSVA to adjust for RNA degradation. This method, which is based on directly measured experimental degradation of RNA in postmortem brain, substantially attenuates the differences observed between LIV and PM tissues. However, RNA degradation is not the only salient difference between the LIV and PM tissues investigated by Liharska et al. In our Discussion, we point out other substantive differences between these two sets of samples, which make this uncontrolled experiment difficult to interpret. We also compare summary statistics from Liharska to those from another postmortem sample. Perhaps surprisingly, these results highlight age-of-death (AoD) and RNA quality as more of a problem for the LIV than the PM tissues; these are likely key differences between the two sets of samples, and together with RNA degradation issues, challenge Liharska’s conclusion (1).

Regarding (2), the results in Liharska actually have no bearing on whether molecular signatures of disease processes can be identified in postmortem samples, which has been demonstrated – and repeatedly replicated – in multiple published studies. Well-designed post-mortem experiments either match samples of affected and unaffected subjects for PMI, RIN, sex, age and other metrics related to RNA degradation or contain enough samples to control, statistically, for effects of these and other covariates. Both have proven replicable, given sufficient sample size; even in Liharska, their contrasts of PM case-control cohorts show congruent results across cohorts. Furthermore, gene expression and splicing signatures from postmortem human brain tissues are significantly heritable in the population and exhibit substantial local genetic correlations with brain disorders (Fromer et al. 2016). Many genetically predicted findings in disease are also found to be differential expressed in postmortem case-control comparisons (Gandal et al. 2018; Raj et al. 2018). If molecular signatures in postmortem brains were truly inaccurate, and not relevant for understanding disease processes, it is hard to imagine why they would be reproducibly associated with germline genetic variation in disease-relevant differentially expressed genomic loci.

It is also worth noting that, in general, comparing LIV to PM from different subjects as done by Liharska et al will be severely confounded with many technical and biological variables. These confounding factors are not always obvious/known, and they are more difficult to control in such LIV to PM design than comparing LIV to LIV, PM to PM. Comparing LIV to PM from the same subjects will be helpful, but it will be difficult to get samples. Sadly, the Liharska data does not even have controls for the case-control DEG analysis in LIV samples, essentially making LIV-only design practically infeasible for most of the brain diseases that we aim to study.

As described in Results and Discussion below, the Liharska LIV versus PM experiment has many uncontrolled confounders. Most notably, the living samples were all derived from individuals undergoing neurosurgical procedures, and exposed to anesthesia, neither of which are the case for the PM tissues being contrast. No statistical correction, however powerful, can ever fully correct for such an exposure perfectly confounded with an outcome of interest. Unfortunately, the many correlation-based analyses that they assert tests for effects of confounders do not remove the effects of the confounders, making their correlation results meaningless. We elaborate this point in what follows.

## Results

### Motivation

The DE patterns observed in Liharska are consistent with a different normalization for LIV versus PM transcriptomes. What might drive such a difference? There are many possibilities. A controlled experiment, however, provides one possible driver among the many. In their 2005 work, entitled “A microarray study of post-mortem mRNA degradation in mouse brain tissue”, Catts et al. (2005) evaluated rates of degradation of gene transcripts as a function of PMI from mouse brain. They raised 40 Balb/c mice, balanced for males and females, and quantified transcript abundance in brain for 5-6 mice at 6 hours intervals, ranging from 0 to 48 hours PMI. While total RNA abundance did not change significantly during the first 24 hours, it dropped by 16% during the next 24 hours. Roughly 10% of transcripts show substantial degradation as PMI progressed from 0 to 48 hours. A similar study of proteomic data from 5xFAD mice had a similar conclusion (Bai et al., 2020).

If Catts et al. had quantified transcripts using RNA-seq, instead of microarrays, it is reasonable to expect their results regarding degradation would be similar. However, when 10% of transcripts substantially degrade, the RNA-seq reads that would have targeted those missing (degraded) transcripts go somewhere else: they are distributed to the 90% of transcripts with little or no degradation, at about their relative frequencies. All other things being equal, about 10% of transcripts would be stochastically lower than expectation and 90% would be higher. This process yields different normalizations for the two “experimental” conditions, LIV and PM. Our simulation experiment mimics this situation, using results from a more recent transcriptomic experiment.

### Simulation experiment

We base our simulations on the gene expression dataset in prefrontal cortex from the CommonMind Consortium (CMC), a dataset described in detail previously (Fromer et al. 2016, Hoffman et al. 2022) and analyzed by Liharska. In the CMC dataset, expression of 16,432 protein coding genes were characterized by RNA-seq. In our analyses of these data, PMI explained significant variation in expression for 4,041 genes (FDR < 0.05), 1,384 (8.4%) decreasing with PMI and 2,657 (16.2%) increasing. We take the estimated distribution of CPM from 258 of the 279 control samples to simulate our LIV sample. In addition, we use the estimated fold change for PMI, distribution of PMIs, and the distribution of the total library size in the control portion of the CMC dataset for our simulations.

To simulate expression counts for LIV sample i, we use a multinomial distribution (function rmultinom in R) with the library size for sample i as the total count and the probability of a count being assigned to gene k being proportional to the average expression for gene k compared to the total average expression. When simulating expression counts for PM sample j, we first adjust the mean expression for each of the genes by the effect of PMI, b_k(PMI)_, estimated from the CMC data and following two scenarios described below. These adjusted mean expressions are then used in the multinomial distribution with total count equal to the library size of sample j. DE between LIV and DN samples are then computed using log_2_(cpm(expression+0.5)) as the outcome and PM (yes=1, no=0) as the predictor using function lm in R. Correction for multiple testing is performed using function qvalue in R. We simulate 258 PM samples using the same library sizes as used for the LIV samples. One hundred data sets are generated and analyzed per scenario.

- Scenario 1 reduces the average expression for the 1,384 genes with q < 0.05 and b_k(PMI)_ < 0 in the CMC data. Call these PMI-down genes. We then proportionately adjust the mean expression of the remaining genes (N=15,048) upwards to account for the expected deficit of reads from the PMI-down genes and the total library size for each sample.
- Scenario 2 reduces the average expression for all 6,306 genes with b_k(PMI)_ < 0, regardless of q-value. The mean expression of the remaining genes is adjusted as in Scenario 1.

Consistent with expectation, in Scenario 1 (**Table 1**) we observe a mean of 1,426.6 PMI-down genes over the 100 experiments (8.7%, FDR < 0.05). Notably, however, there are now 5,929.9 PMI-up genes, on average, where none was expected if we were measuring abundance on an absolute scale. Moreover, 44.8% of genes show significant DE, whereas only 8.4% were set to PMI-down. Results from Scenario 2 are like Scenario 1 (**Table 1**), although more extreme: on average, 12,701.7 genes are DE (77.3%), with roughly 3,919 PMI-down and 8,783 PMI-up. The proportion being DE genes, 77.3%, is remarkably similar to that found by Liharska for their LIV-PM contrast, 79.4%. The ratio of the counts of PMI-up and PMI-down genes is not similar, however, showing that RNA degradation due to PMI cannot be the sole driver of Liharska’s LIV-PM DE results.

**Table 1.** Number of differentially expressed (DE) genes between simulated LIV and PM conditions.

| Model | q < 0.05 | Scenario 1 |  | Scenario 2 |  |
| --- | --- | --- | --- | --- | --- |
|  |  | Mean | StDev | Mean | StDev |
| DE | ALL | 7365.5 | 89.5 | 12,701.7 | 105.3 |
|  | DOWN | 1426.6 | 6.7 | 3919.0 | 29.1 |
|  | UP | 5929.9 | 86.5 | 8782.7 | 82.3 |
| DE + PMI | ALL | 0.1 | 0.2 | 0.1 | 0.4 |
|  | DOWN | 0.0 | 0.2 | 0.0 | 0.1 |
|  | & UP | 0.0 | 0.2 | 0.1 | 0.3 |

In Liharska, the LIV samples have a PMI of zero, approximately, whereas the PM samples have a positive PMI. This is true for our simulation scenarios as well. We wondered if we fit PMI to the simulated data, including the LIV samples, would this modeling adjust appropriately? Regardless of scenario, fitting PMI to the data yields virtually no DE at FDR < 0.05 (**Table 1**), consistent with the simulated truth.

### Re-analysis of Liharska adjusting for degradation

We downloaded the FASTQ files from Synapse (syn26337520) and aligned them against GRCh38 and Gencode v25 using SPEAQeasy (Eagles et al. 2021). After merging the sample phenotypic data provided by Liharska et al, we explored the association between different RNA sequencing and alignment quality metrics provided by SPEAQeasy against the top 10 principal components (PCs, **Fig 1. A, Table S1**). This showed an association between PC1 (27.6% of variance explained) and the postmortem (LIV vs PM) tissue status, but also between PC1 and other covariates such as RNA integrity number (RIN) and the fraction of mapped reads assigned to genes (totalAssignedGene). Postmortem tissue status was more strongly associated with PC2 (6.2%) and PC3 (4.6%), with PC2 being associated with totalAssignedGene, rnRNA rate, RIN, age at time of death and PC3 associated with the proportion of mitochondrial reads (mitoRate), totalAssignedGene, RIN, rRNA rate (**Fig 1. A**). LIV and PM tissues showed significant differences in some of these quality metrics including mitoRate and totalAssignedGene (**Fig 1. B, Table S2**), with both being relatively more problematic in the LIV than the PM tissue. LIV and PM tissues also showed significant differences in the estimated proportions of oligodendrocytes and neurons (**Fig 1. C, Table S3**), as revealed by estimating cell type proportions using data from Darmanis and colleagues as the reference (Darmanis et al. 2015, Collado-Torres et al. 2019). To generate a new variable from these deconvolutions, we summarized the estimated cell type proportions across the ten cell types in Darmanis et al. through principal component analysis and extracted the first PC (79% of variance explained), which we refer to as cellPC1. Next, using the SPEAQeasy aligned data, we applied the limma-voom differential expression framework (Ritchie et al. 2015) to re-calculate gene level t-statistics between LIV and PM tissues, after adjusting for race, sex, diagnosis, RIN, cellPC1, mitoRate, totalAssignedGene, rRNA_rate, and overallMapRate. This analysis results in 15,553 differentially expressed genes (FDR < 0.05), although the range of these re-calculated t-statistics was smaller and overall attenuated compared to the original results by Liharska et al. (**Fig 1. D, Table S4**). Using bulk RNA-seq data from a brain tissue degradation experiment on the dorsolateral prefrontal cortex (Jaffe et al. 2017), processed with the recount3 pipeline (Wilks et al. 2021), we contrasted at the transcript level the t-statistics for degradation against t-statistics for LIV vs PM tissue differences that we re-calculated using limma (Ritchie et al. 2015), resulting in a correlation of 0.39 (**Fig 1. E, Table S5**). This correlation reveals confounding between the LM vs PM tissue differences and RNA degradation. To adjust for RNA degradation, we generated 16 quality surrogate variables (qSVs, Jaffe et al. 2017) from the 2,924 transcripts that have been shown to be significantly associated with RNA degradation, while also adjusting for cell proportion differences (Stolz et al. 2023) that were observed in the Liharska et al. dataset. At the gene level, we observed an attenuation of the LIV vs PM tissue differences when adjusting for the 16 qSVs (**Fig 1. F, Table S4**) resulting in 3,732 differentially expressed genes (FDR < 0.05), a 75% reduction compared with DE without qSVA adjustment.

**Figure 1.**
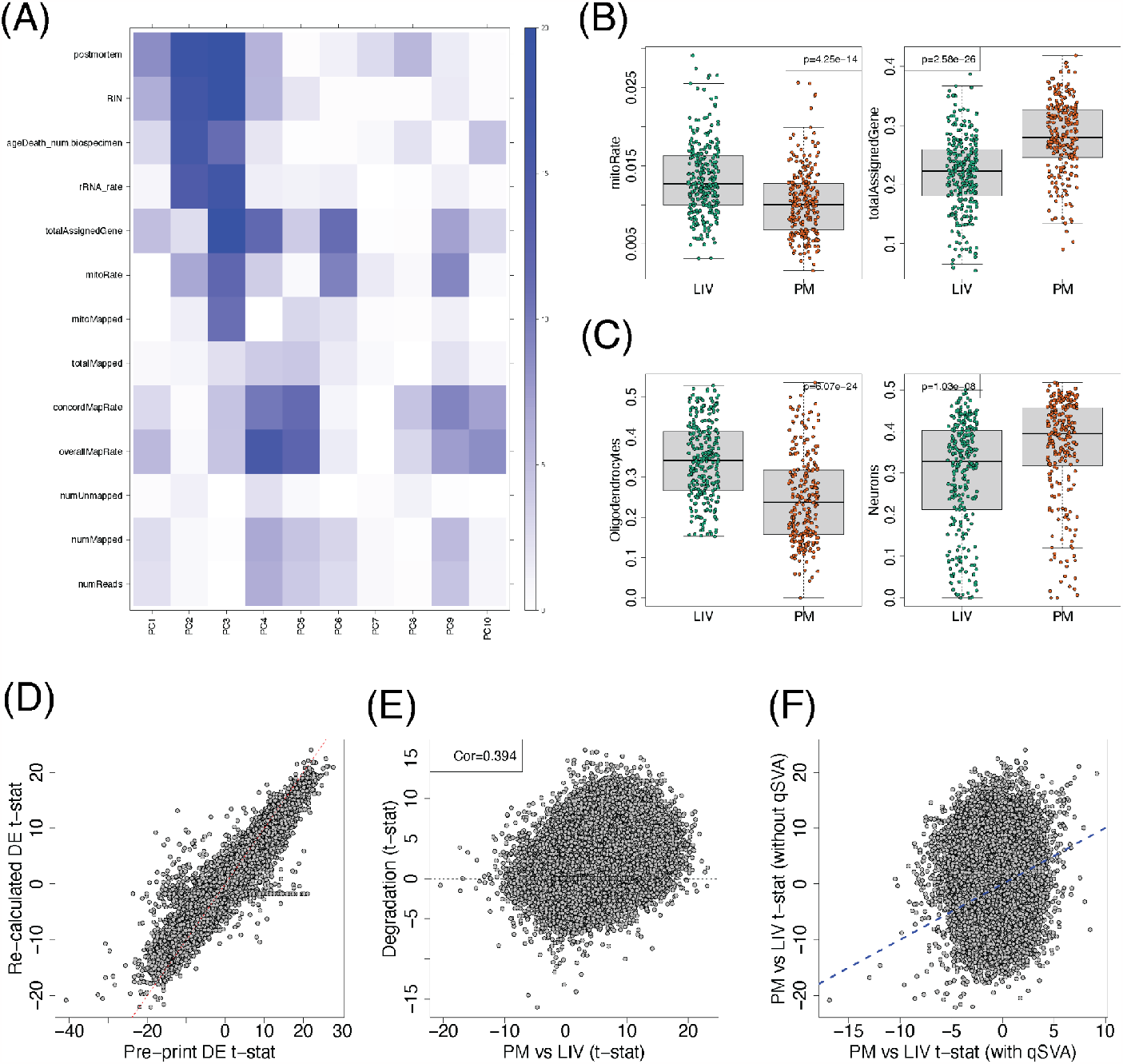
Re-analysis of the Lisharka dataset. (**A**) Heatmap of the -log10(p-value) for the correlation between RNA-seq quality metrics, RIN, age at time of death, and postmortem status (LIV or PM) against the top 10 principal components (PCs, **Table S1**). (**B**) Mitochondrial mapping rate (mitoRate) and the proportion of mapped reads assigned to genes (totalAssignedGene) are quality metrics that are significantly different between LIV and PM samples (two-sided t-test, **Table S2**). (**C**) Estimated proportions for oligodendrocytes and neurons are significantly different between LIV and PM samples (two-sided t-test, **Table S3**). (**D**) Gene level comparison of the LIV vs PM t-statistics calculated by Liharska et al. (pre-print) against our re-calculated results using the SPEAQeasy alignments and the limma-voom framework adjusting for race, sex, diagnosis, RIN, cellPC1, mitoRate, totalAssignedGene, rRNA_rate, and overallMapRate (**Table S4**). (**E**) Transcript level DEqual plot comparing LIV vs PM t-statistics from the same model against degradation t-statistics from the Jaffe et al. 2017 degradation data (**Table S5**). (**F**) Gene level PM vs LIV t-statistics contrasting a model with and without adjusting for quality surrogate variables (qSVs, **Table S4**).

## Discussion

Finding genes showing differential expression (DE) between LIV and PM tissue is unsurprising. The issue is what drives the differences and what you can conclude from them. The many differences between the LIV and PM brain tissues reported by the Liharska study make comparisons problematic. (Even comparisons of separate postmortem samples can be challenging.) Liharska’s analyses have not adequately controlled for PM interval in PM tissue, let alone RNA degradation, which is not adequately represented only by PMI (Jaffe et al. 2017).

A number of factors showed surprising difference between LIV and PM tissues, including the RNA Integrity Numbers (RIN) and proportions of multiple cell types. RINs of the LIV tissue from Liharska are surprisingly low; on average, they are lower than the RIN on the PM tissue. This is odd because fresh tissue should not show such evidence of poor RNA quality, and some studies have found a connection between neurodegeneration and lower RIN (Highet et al. 2021 and references therein). The differences in proportion of neurons in the two tissue datasets is also a potential unexplained biological confounder, and of even greater concern is the dramatic variance in neuronal proportion in the LIV compared to the PM samples. In fact, over 50% of the LIV samples have neuronal proportions below the 75th percentile of that from PM samples. In our experience, such a striking discrepancy in neuronal proportion is often a result of systematic differences in tissue dissection and experimental handling. For example, careful identification of the gray-white matter boundary within frozen PM tissue enriches for neuronal populations rather than oligodendrocytes. However, neurosurgical biopsy in LIV samples may not allow such careful dissection, which may explain the substantial increase in non-neuronal cell-types, such as oligodendrocytes. Another possibility is that there is much more gliosis in the LIV samples, since they are largely derived from individuals with neurodegeneration. In any event, Liharska assumes that accounting for neuronal fraction in their models can adequately account for the immense diversity of cells in the human brain. However, given the differences between LIV and PM tissue for Liharska’s binary representation (neuron/other), it is unreasonable to expect this predictor can account for the underlying diversity of cell types, some of which likely show just as much heterogeneity as the binary representation itself. In our re-analysis we used PCA on the estimated cell type proportions and retained cellPC1 to control for variation in cell-type representation; alternative modeling would include either using more cell-based PCs as covariates or a method like isometric log ratio, as in other studies (Hoffman et al 2022).

Our simulation experiments show how ignoring RNA degradation, here due solely to PMI, can lead to vast DE differences between LIV and PM tissue. The DE extends beyond the genes with degraded RNA. Even when the absolute abundance of RNA transcripts from most genes are not different between LIV and PM tissue, the relative nature of RNA-seq generates substantial DE. Even so, it is likely that RNA degradation is a greater driver in DE because the LIV tissue shows lower RIN, higher mitochondrial mapping and lower gene mapping rates. Importantly, this is just one of the many differences between the LIV and PM tissue.

To explore what might be driving the differences, we first asked the authors for their summary statistics for the LIV-PM contrast, from which we extract the estimated t-statistic for each gene. We next fit the CMC data to a set of covariates used in Fromer et al., which were shown to be associated with expression for subsets of genes. These were PMI, age-at-death (AoD), RIN, and RIN^2^, and they were fitted jointly to expression of each gene. The estimated t-statistic for each of these variables was then obtained. Being two different data sets, LIV-PM versus CMC, the expected correlations between the LIV-PM estimated *t*-statistic and those from the CMC’s covariates would be zero, unless the LIV-PM analysis did not adequately control for the effects of the covariates. In fact, we found notable correlations for all covariates (**Fig. 2**), especially for AoD and RNA quality (RIN and RIN^2^).

**Figure 2.**
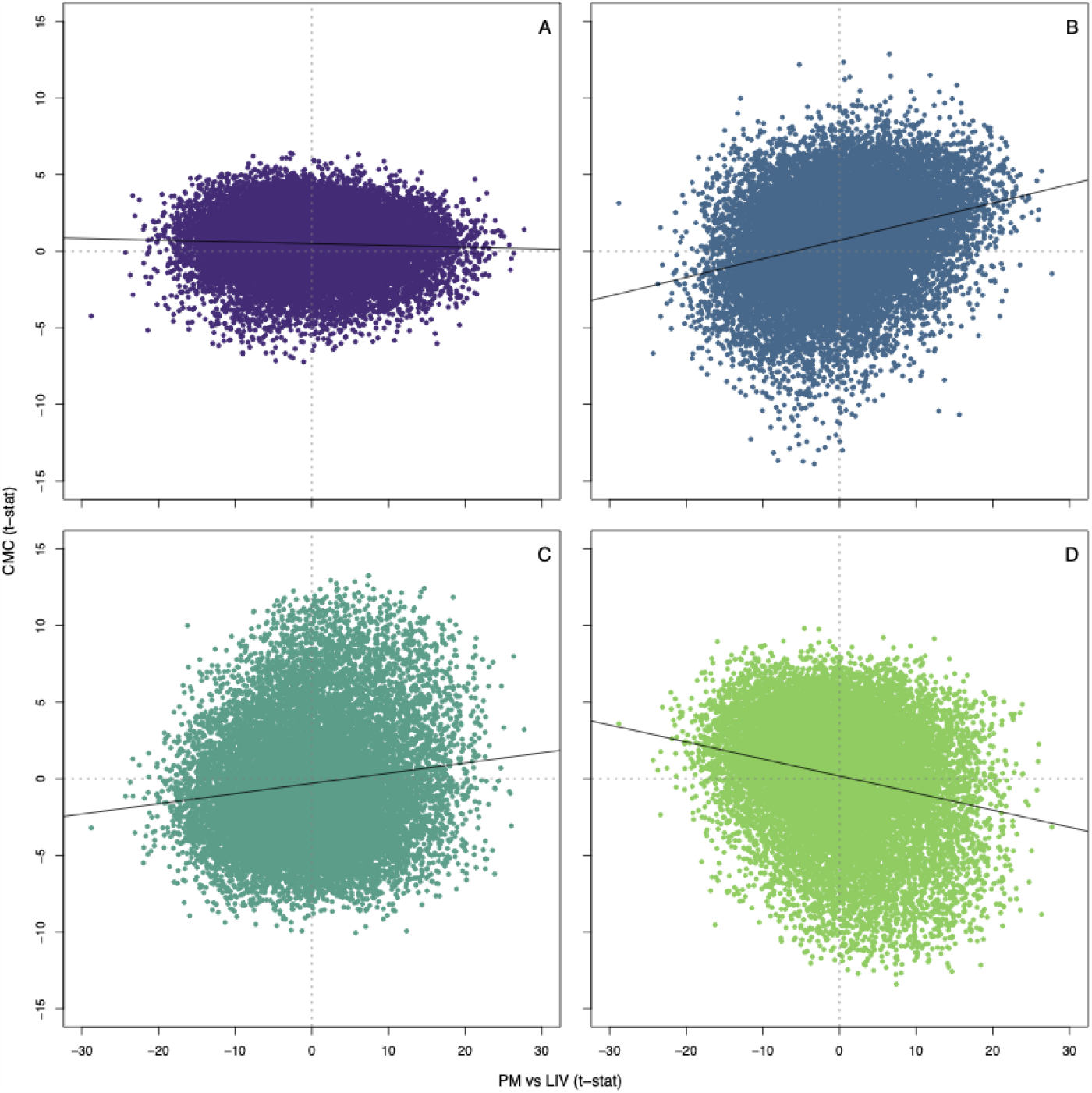
Effects of LIV-PM versus effects of covariates in the CommonMind Consortium study. (Fromer et al. 2016). The test-statistic for the contrast of PM versus LIV tissue, PM vs LIV(t-stat), is plotted against the test statistics for covariates included in the modeling of the CommonMind (CMC) data, CMC (t-stat). Covariates included in the Fromer et al. study were (**A**) Post-mortem interval, PMI; (**B**) age-of-death, AoD; (**C**) RNA integrity number, RIN; and (**D**) the square of RIN. Correlation r (p-value) for PMI, r = -.046 (1.12 × 10-8); for AoD, r = .282 (3.02 × 10-220); for RIN, r = .133 (8.86 × 10-62); for the square of RIN, r = -.227 (9.33 × 10-180).

These results prompted us to obtain the Liharska et al. dataset so that we could delve more deeply into LIV-PM differences. By re-aligning the Liharska dataset and re-analyzing their data, we showed that, while LIV vs PM tissue differences are prominent, there are other RNA sequencing quality metrics that were strongly associated with the top principal components (**Fig. 1 A**). Exploratory results showed that there were significant differences between the LIV and PM tissue on these quality metrics (**Fig. 1 B**), with surprisingly lower quality on the LIV tissue compared to the PM tissue. We also showed how we can mostly reproduce their statistical model (**Fig. 1 D**), prior to highlighting how the LIV vs PM differences are confounded with RNA degradation signal observed independently (Jaffe et al. 2017, Fig. 1 E). Using the latest quality surrogate variable method, we showed how the differences between LIV and PM tissues were substantially attenuated, resulting in a reduction of 76% in the number of differentially expressed genes (15,553 vs 3,732). These results highlight that RNA degradation is a crucial confounder in the differences between LIV and PM tissue observed by Liharska et al. Furthermore, the degradation effects can be addressed analytically by methods like qSVA. Given these observations, we believe that the conclusions by Liharska et al about the differences between LIV and PM tissue need to be reassessed.

In an attempt to validate their results, Liharska undertakes a series of correlation analyses, each of which correlates the original LIV versus PM DE results with some subset of the data. Apparently, their expectation is that the correlation would tend toward zero if the original results were invalid. Because their correlations always approach one, they believe the results are valid. Our simulation experiment (**Table 1**) and contrast with CMC covariate effects (**Fig. 2**) reveal the flaw in this logic. Each new contrast has the same underpinning weaknesses, such as RNA quality, degradation, and AoD. If one does not remove the effect of important confounders, they will drive the correlation results. Their subsets involving illness, treatment, and anesthesia in the LIV cohort all have this same weakness. If it were possible to remove the effects of all confounders from both the LIV and PM cohorts, and only then evaluate DE, that would be a compelling evaluation of how different the LIV and PM transcriptomes are.

There are orthogonal approaches to evaluate the nature and quality of the LIV and PM data and whether they yield similar information regarding transcriptomic effects from age, disease, and genetic variation. If the LIV and PM samples were similarly informative, we would expect similar effects of age, phenotype, and genotype from these data sets. For example, one might identify eQTLs (expression Quantitative Trait Loci) from each dataset and calculate what fraction of eQTLs they share. When this was done for LIV (Young et al., 2021) versus PM microglia (Lopes et al. 2022), the estimated fraction of eQTL shared was uniformly high (see Fig. 4d in Lopes et al. 2022). One might also ask if the well-known effects of age on gene expression is reflected in both data sets. Curiously, according to their Methods, Liharska did not include age as a covariate in their models, despite age being different between the cohorts, though they did explore the interaction of age with disease status, absent a main effect of age. Although in our re-analysis we note that age at time of death is equally associated with PC2 and PC3 (**Fig. 1 A**), we did not adjust for it in our re-analyses in an effort to focus on the effect of RNA degradation (**Fig. 1 D-F**). As noted above, one wants to estimate the effects of age and remove it – as well as those of important confounders in these two data sets – before computing DE signatures and comparing them. Liharska assert that they cannot remove the effect of PMI because the LIV samples have no value for PMI. We disagree, their PMI value is essentially zero. Indeed, as we show in our analyses, modeling PMI removes spurious DE from the LIV-PM contrast in our simulations (**Table 1**). One might also estimate DE associated with Parkinson’s disease in each dataset. However, this is more challenging for the LIV tissue, which contains 220 samples from patients diagnosed with Parkinson disease and 55 samples from patients with another diagnosis. Most of the 55 subjects were diagnosed with dystonia or essential tremor, neither of which is etiologically distinct from Parkinson disease (Thenganatt and Jankovic 2016; Shetty et al. 2019). Teasing out associations specific to Parkinson disease could be compromised by these confounding disease effects in the patients without a Parkinson diagnosis.

The difficulty of data sets having different scaling or normalization is not new, and there are methods available to make such datasets more similar. Peng and colleagues (2021), for instance, introduce cFIT, for common factor integration and transfer learning, to model batch effects across experiments (e.g., LIV versus PM). cFIT assumes there is shared information between datasets that can be captured by a common factor space, even when expressions of genes show distortions specific to batch.

We conclude with a comment about whether gene expression data from postmortem brain tissue has yielded important insights into the genetic mechanisms of brain disorders. Across the spectrum of developmental disorders (eg. ASD, schizophrenia) to neurodegenerative disorders (e.g Alzheimer’s Disease, Parkinson’s disease) the gene expression data have translated clinical genetics into credible and functional pathogenic mechanisms of illness that have been confirmed in cell and animal model systems and elucidated cell types of primary pathogenic relevance, none of which was known without these studies. Analyzing transcriptome data requires proper evaluation and control of covariates. Otherwise, it could lead to enormous false findings. Moreover, it is worth noting that even if none of the confounders and limitations noted above were at issue, living tissue, which is available only under anesthesia and in most instances from a small section of prefrontal cortex in older individuals with chronic brain illness, is a markedly limited resource, and far from a gold standard in studying the spatially and developmentally complex molecular mechanisms of brain illness. It is unethical to collect brain tissues from most living people, particularly healthy. That makes the studying living tissue useless for at least case-control design. Living tissues will be useful for studies of genetic and environmental factors with supposedly better quality of RNA and little postmortem effects.

## Data Availability

All re-analysis results, re-processed LBP and qSVA datasets, and original datasets are available online at

https://github.com/LieberInstitute/living_brain_reanalysis

https://adknowledgeportal.synapse.org/Explore/Studies/DetailsPage?Study=syn26337520

https://www.synapse.org/CMC

https://doi.org/10.7303/syn23763487

https://www.ncbi.nlm.nih.gov/Traces/study/?acc=SRP108559

## Acknowledgements

We would like to thank Andrew E. Jaffe for conceptualization of and help with the re-analysis of the Liharska et al. data. The results published here are in whole or in part based on data obtained from the AD Knowledge Portal (https://adknowledgeportal.org/). The Living Brain Project study participants are commended for their important role in science. The Living Brain Project is funded by the National Institute of Aging (R01AG069976) and the Michael J. Fox Foundation (Grant Number 18232). Postmortem samples from Harvard Brain Tissue Resource Center and University of Miami Brain Endowment Brain were acquired under the National Institutes of Health NeuroBioBank request number 543. Postmortem samples from the New York Brain Bank of Columbia University were acquired under request number 1962. CommonMind Consortium data was utilized under the National Institutes of Mental Health Repository and Genomics Resource request identifier 5bffe32e97024. We thank the original authors for their prompt responsiveness and for sharing their data, which allowed us to perform this re-analysis.

## Code and Data Availability

The LBP study data is available from Synapse (syn26337520): https://adknowledgeportal.synapse.org/Explore/Studies/DetailsPage/StudyDetails?Study=syn26337520. The CMC data is available from https://www.synapse.org/CMC and https://doi.org/10.7303/syn23763487. The qSVA data (Jaffe et al. 2017) is available from https://www.ncbi.nlm.nih.gov/Traces/study/?acc=SRP108559. Our re-analysis code and main *SPEAQeasy* re-processed LBP and qSVA data are available at https://github.com/LieberInstitute/living_brain_reanalysis (Collado-Torres 2023).

## Supplementary Material

### Supplementary Tables

**Table S1**: Table with the p-values for the correlation between the top 10 gene-level principal components (PCs) and several *SPEAQeasy* metrics as well as the LIV vs PM indicator variable. The -log_10_ p-values are displayed in **Figure 1A**.

**Table S2**: Merged sequencing metrics produced by *SPEAQeasy*; see https://research.libd.org/SPEAQeasy/outputs.html#quality-metrics for details about the *SPEAQeasy* alignment metrics. Sample IDs are the same ones used by Liharska et al and were not known to anyone outside the research group. Data from this table is used for **Figure 1B**.

**Table S3**: Per sample, estimated cell type proportions via deconvolution. Sample IDs are the same ones used by Liharska et al and were not known to anyone outside the research group. Data from this table is used for **Figure 1C**.

**Table S4**: Limma-voom gene-level differential expression results between LIV and PM samples either (a) without adjusting for qSVs (quality Surrogate Variables), (b) adjusting for qSVs, or (c) reproducing the original results by Liharska et al. Data from this table is used for **Figure 1D** and **Figure 1F**.

**Table S5**: Limma transcript-level differential expression results between (a) LIV and PM samples or (b) by RNA degradation (Jaffe et al., *PNAS*, 2017). Data from this table is used for **Figure 1E**.

